# Duration of viral detection in throat and rectum of a patient with COVID-19

**DOI:** 10.1101/2020.03.07.20032052

**Authors:** Le Van Tan, Nghiem My Ngoc, Bui Thi Ton That, Le Thi Tam Uyen, Nguyen Thi Thu Hong, Nguyen Thi Phuong Dung, Le Nguyen Truc Nhu, Tran Tan Thanh, Dinh Nguyen Huy Man, Nguyen Thanh Phong, Tran Tinh Hien, Nguyen Thanh Truong, Guy Thwaites, Nguyen Van Vinh Chau

**Author notes:** **Correspondence** Le Van Tan, Oxford University Clinical Research Unit, Ho Chi Minh City, Vietnam, Nguyen Van Vinh Chau, Hospital for Tropical Diseases, Ho Chi Minh City, Vietnam.

## Abstract

The rapid spread of coronavirus disease 2019 (COVID-19) raises concern about a global pandemic. Knowledge about the duration of viral shedding remains important for patient management and infection control. We report the duration of viral detection in throat and rectum of a COVID-19 patient treated at the Hospital for Tropical Diseases in Ho Chi Minh City, Vietnam. Despite clinical recovery, SARS-CoV-2 RNA remained detectable by real time RT-PCR in throat and rectal swabs until day 11 and 18 of hospitalization, respectively. Because live SARS-CoV-2 has been successfully isolated from a stool sample from a COVID-19 patient in China, the results demonstrate that COVID-19 patients may remain infectious for long periods, and fecal-oral transmission may be possible. Therefore, our finding has important implications for infection control.

---

The emergence of severe acute respiratory syndrome coronavirus 2 (SARS-CoV-2) and its current global spread have prompted the World Health Organization to declare a Public Health Emergency of International Concern. We report the duration of viral detection in throat and rectal swabs taken from a patient treated at the Hospital for Tropical Diseases (HTD) in Ho Chi Minh City, Vietnam. HTD is a referral hospital responsible for receiving and treating patients with coronavirus disease 2019 (COVID-19) from southern Vietnam with a population of >40 million.

On January 31, 2020, a 73-year-old man with a history of benign prostatic hyperplasia was admitted to HTD with dry cough and breathing difficulties, but without fever. He had become ill with respiratory symptoms on January 26, 2020, ten days after arrival in Vietnam from the United States. Whilst travelling to Vietnam, he had two hours of transit at an airport in Wuhan, China.

Blood tests on hospital admission showed an elevation of white cells (22,9×10^3^ cells per cubit millimeter), but others parameters (platelet and hematocrit) were in normal ranges (data not shown). His chest radiographs obtained on admission showed bilateral infiltrates (Supplementary Figure 1). The patient was empirically treated with a combination of antivirals and antibiotics, and received supplemental oxygen through an oxygen mask.

On February 1, 2020, a throat swab collected on admission from the patient was positive for SARS-CoV-2 by reverse-transcription polymerase chain reaction (RT-PCR) (Supplementary Table 1), followed by sequencing of the amplified product to confirm the result. Testing for other pathogens (routine bacterial culture and influenza A/B viruses) were negative.

His clinical condition improved, and from February 9^th^ he no longer required supplementary oxygen. However, follow-up throat swabs remained positive for SARS-CoV-2 by real-time RT-PCR (1), until day 11 of hospitalization (i.e. day 16 of illness) (Figure 1). Rectal swabs were RT-PCR positive until day 18 of hospitalization (day 23 of illness). Additionally, a plasma sample collected on day 2 of admission was weakly positive; urine samples were negative (Figure 1). The patient was discharged with full recovery on February 21, 2020 after 21 days of hospitalization.

**Figure 1:**
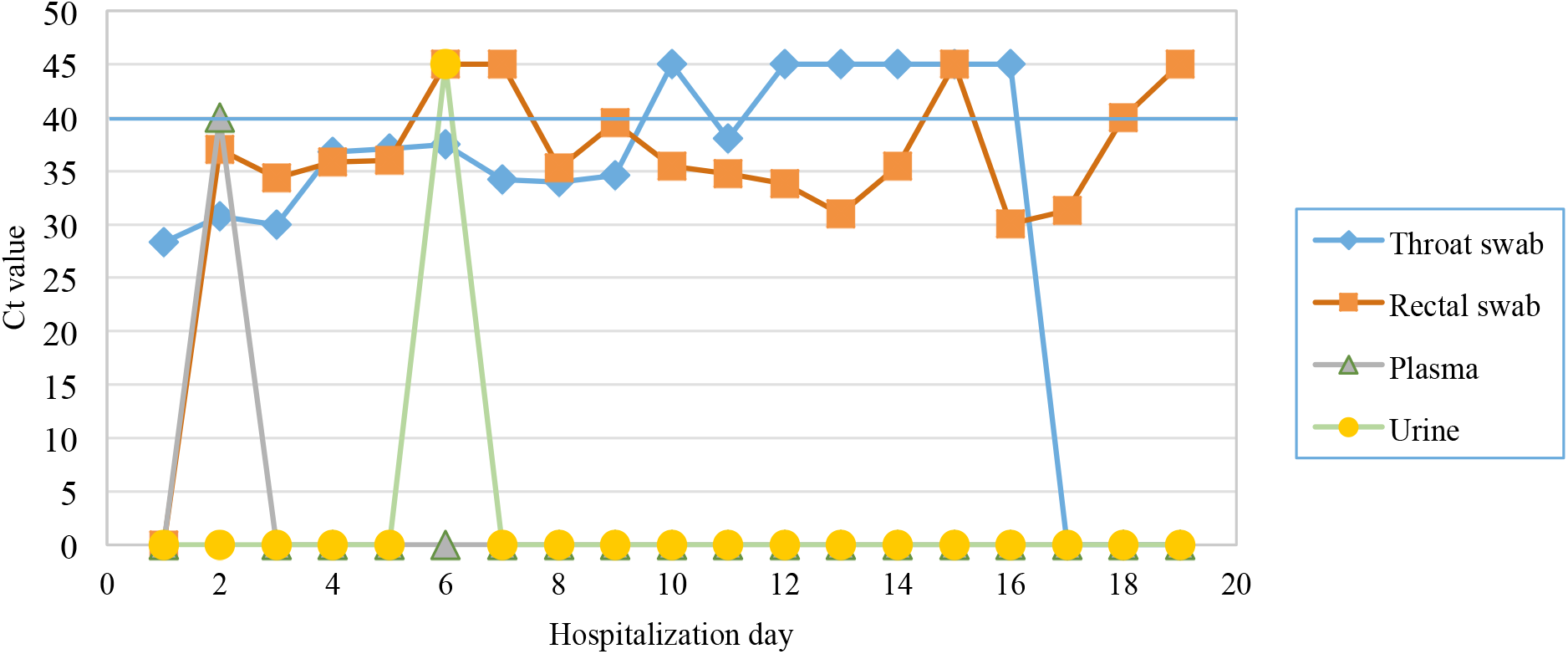
Results of real-time RT-PCR analysis of serial samples **Note to figure 1:** a Ct value of 0 indicates no samples available for RT-PCR analysis. Ct value of 40 is the cut-off of assay detection limit (blue line). A Ct value of 45 represents for a negative real-time RT-PCR result.

Since his arrival in Vietnam, the patient had stayed in a hotel in Ho Chi Minh City, and had sought for medical treatments at private clinics before hospital admission. A total of 14 close contacts were identified and followed for 14 days, but none had signs of respiratory infections, and their throat swabs collected on day 14 of quarantine were RT-PCR negative for SARS-CoV-2.

Long duration of viral detection in throat swabs by RT-PCR has been reported in a case series from China (2) and a patient in the US (3). Together with the recent success of SARS-CoV-2 isolation in cell culture in a stool sample of a COVID-19 patient in China (4), our finding on the persistent PCR positivity in rectal swabs raise concerns about the possibility of fecal-oral transmission of SARS-CoV-2.

Collectively, our report shows the persistence of PCR positivity in throat and rectum of a patient with COVID-19, which may potentially indicate persistent viral shedding. As such COVID-19 patients may remain infectious for long periods, and fecal-oral transmission may be possible. Therefore, our finding has important implications for infection control.

## Data Availability

All data referred to in the manuscript have been presented either in the manuscript main text or in Supplementary Materials

## Acknowledgements

This study was funded by the Wellcome Trust of Great Britain (106680/B/14/Z and 204904/Z/16/Z).

We thank Pham Thi Ngoc Thoa, Tran Nguyen Phuong Thao, Tran Thi Lan Phuong, Tran Thi Thanh Tam, Huynh Thi Kim Nhung, Ngo Tan Tai, Vo Trong Vuong, Le Kim Thanh, Le Nguyen Truc Nhu, Nguyen Thi Han Ny, Motiur Rahman and Evelyne Kestelyn for their laboratory/logistic support.

Ware indebted to the patient for his participations in this study, and all the nursing and medical staff at Ward D of the Hospital for Tropical Diseases who provided care for the patient and helped collect clinical data.

## Supplementary Materials

**Supplementary Table 1:**
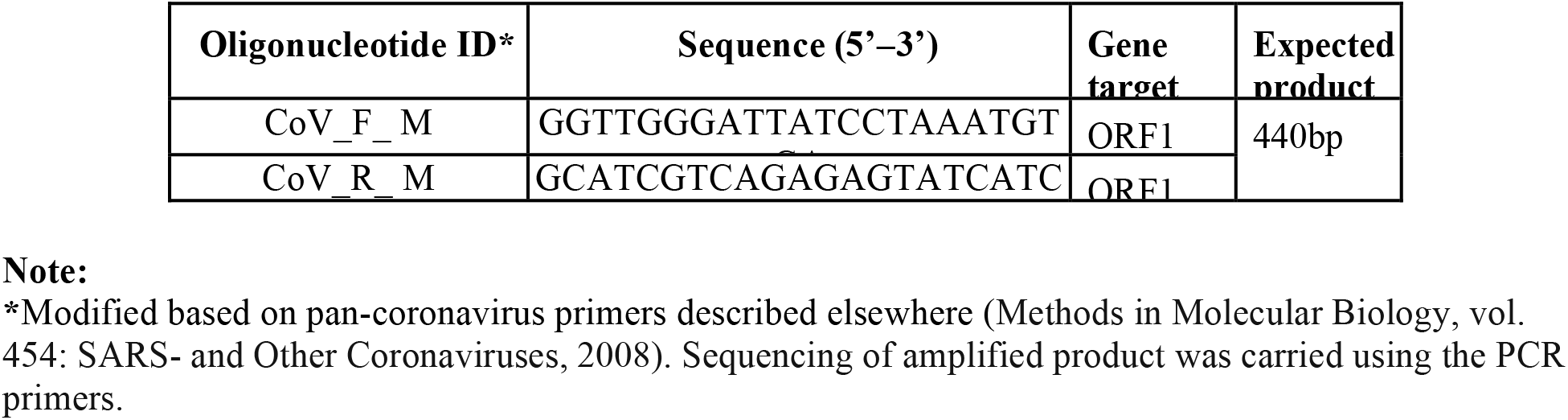
List of oligo sequences used for initial detection of SARS-CoV-2 in admission throat swab of the patient

**Supplementary Figure 1:**
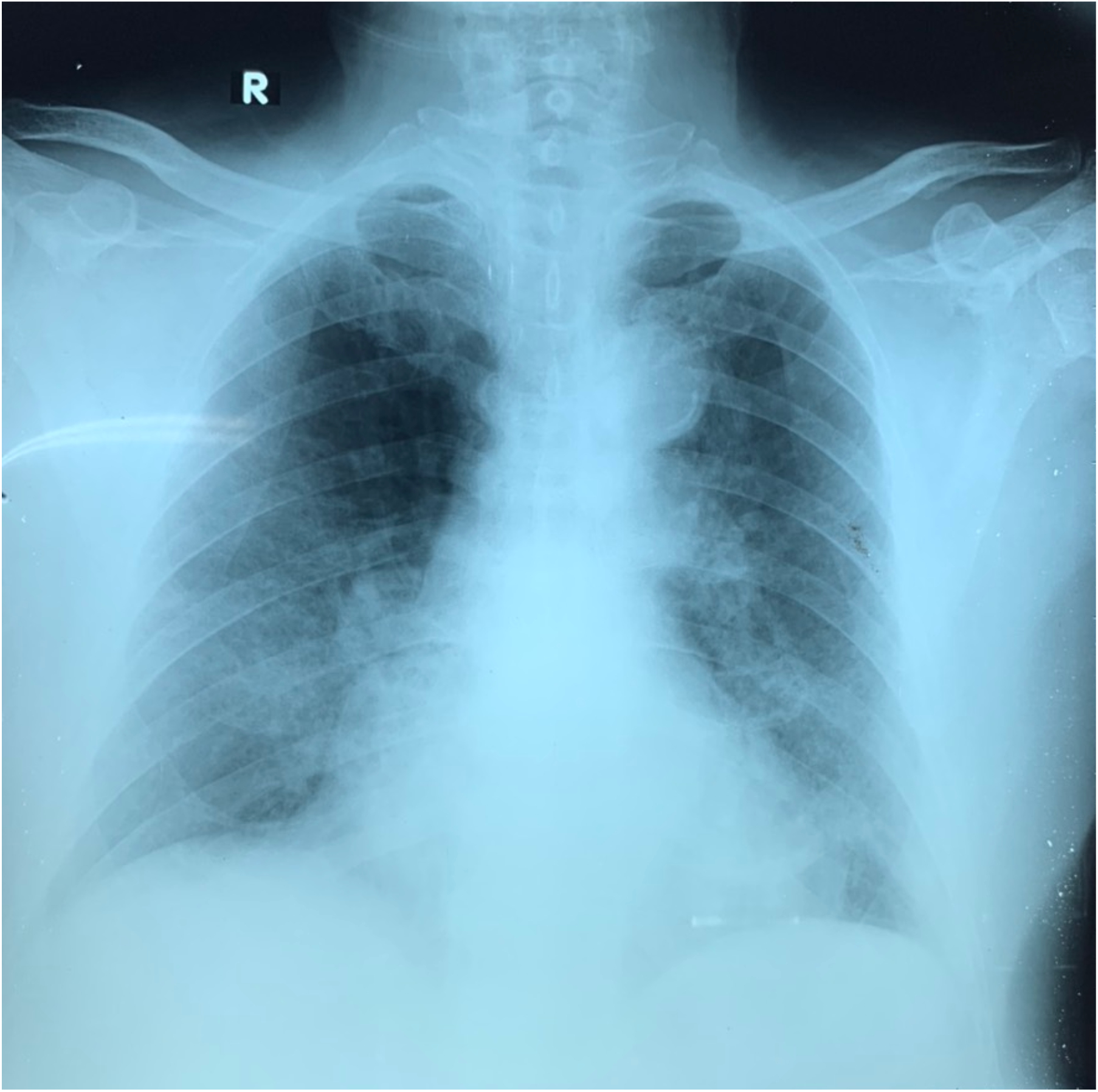
Chest radiograph taken on admission (January 31^st^, 2020) showing bilateral infiltrates

